# SEVERITY AND MORPHOLOGICAL CLASSIFICATION OF ANAEMIA AMONG CHILDREN AGED 2- 59 MONTHS IN DAR ES SALAAM, TANZANIA: A CROSS SECTIONAL STUDY PROTOCOL

**DOI:** 10.1101/2022.11.10.22282169

**Authors:** Salha Ally Omary, Florence Salvatory Kalabamu, Maulid Rashid Fataki, Shani Shamsi Salum, Ummulkheir Hamid Mohamed, Joseph Gasper Kimaro, Kelvin Melkizedeck Leshabari

**Affiliations:** Dept of Paediatrics & Child Health, Hubert Kairuki Memorial University – New Hub street, Dar es Salaam, Tanzania; Dept. of Paediatrics & Child Health, Temeke Regional Referral Hospital – Temeke, Dar es Salaam, Tanzania; Ageing Research group, Registered Trustees of Ultimate Family Healthcare – 11102 Domus Bethania, 12 Olympio Street Upanga – Dar es Salaam, Tanzania; Family Health Research group, Registered Trustees of Ultimate Family Healthcare – 11102 Bethania House, 12 Olympio Street Upanga – Dar es Salaam, Tanzania; Dept of Paediatrics & Child Health, Muhimbili National Hospital (Mloganzila) – Dar es Salaam, Tanzania; Dept. of Paediatrics & Child Health, Msambweni County Referral Hospital - Kwale, Kenya; Dept. of Obstetrics & Gynaecology, Temeke Regional Referral Hospital – Temeke Dar es Salaam, Tanzania

**Keywords:** Anaemia, children, Temeke, Dar es Salaam

## Abstract

**Background:** Anaemia is a clinically significant secondary diagnosis in children under the age of five in most parts of developing world, including Tanzania. Studies that assess clinical conditions associated with prenatal and postnatal era are highly relevant to global health as they tend to reflect patterns of significant health challenges across the entire human lifespan.

**Objective:** To assess for morphological characteristics and severity of anaemia among under-five population seen at Dar es Salaam regional referral facilities in Tanzania.

**Design & Methods:** A cross-sectional hospital based study will be conducted at Temeke Regional Referral and Mbagala Rangi Tatu district hospitals in Dar es Salaam, Tanzania. All children aged 2-59 months will be eligible to participate in the study. All children aged 2 – 59 months with anaemia will be the target population. The study main tool will be a self-administered questionnaire with five distinct parts. Data analysis will commence with summarisation. Specifically, continuous data will be summarised using median (with inter-quartile range) and categorical data will be summarised using frequency (and proportion by %) Graphical tools will also be employed where by important correlations as well as +/- outliers will be assessed. Besides, univariate and bivariate statistics will be computed for all clinically relevant data. Main outcome measure will be severity and morphological classification of anaemia. Unless otherwise stated, an alpha-level of 5% will be used as a limit of type 1 error in findings. Written informed consent will be sought from the parent/guardian of each participant child prior to inclusion into the study.

## 1. Introdution

Anaemia is a clinically significant secondary diagnosis in children under the age of five in most parts of developing world, [1-4] including Tanzania. [5-7] It contributes to various health problems such as recurrent infections, delayed developmental milestones, poor academic performance, and increased morbidity and mortality. [7, 8] Besides, there is mounting evidence that suggest the increased morbidity (anaemia inclusive) in this age group to be a recipe for later years chronic illnesses and premature deaths. [9-11] Thus, studies that assess clinical conditions associated with prenatal and postnatal era are highly relevant to global health as they tend to reflect patterns of significant health challenges across the entire human lifespan. We therefore designed a study to account for morphological characteristics and severity of anaemia among under-five population seen at Dar es Salaam regional referral facilities in Tanzania.

Accordingly, there are conflicting reality on capacity of the health system in developing countries, and especially Tanzania to identify and manage anaemia in children. [12-15] For instance, WHO – SARA 2020 documented that, in Tanzania only 57% of health facilities can test hemoglobin levels among children and 68% of the facilities offered iron supplementation for children, (12) with near-complete lack of resources to diagnose anaemia at an early stage, so anaemia likely went undetected and untreated at dispensaries and health centers. [12] It is therefore prudent to assess attributes associated with anaemia in children in resource limited areas, as that will provide real-world and timely data of the situation at hand.

Reflecting nosology, anaemia can be classified morphologically based on red blood cells and their indices, which makes it easier for health care practitioners to identify and treat anaemia. (16) The morphological classifications of anaemia are normocytic normochromic, microcytic hypochromic, macrocytic, and dimorphic anaemia, with each kind implying different possible causes (17), with the most common type of anaemia being iron deficiency anaemia.(17, 18) Moreover, since anaemia is always a secondary diagnosis, assessment of severity and morphological classification of anaemia is crucial in timely diagnosis and proper management. Despite the Tanzania’s government effort to decrease prevalence and mortality due to anaemia but still, anaemia is a top ten condition with a mortality rate of 9% and also increase the flow of patients among children less than 5 years in both outpatient departments (OPD), and inpatient department (IPD) by 4% and 10% respectively in Temeke municipal council.(19) At present, there is no evidence of the burden and severity of anaemia among children aged 2 to 5 years in Temeke, Dar es Salaam despite the district being the largest of all in Dar es Salaam city.

The World Health Assembly back in 2012, made a resolution 65.6 that endorsed a comprehensive implementation plan on maternal, infant and young children nutrition. (20) The pan specified six different global nutrition plan targets for 2025. (21) Grossly, the plan aims to alleviate the double burden of malnutrition in children, starting from the earliest stages of development. They argued substantial benefits can be guaranteed by concentrating efforts from conception through the first two years of life. (20) Besides, there is hard and solid evidence in literature that supports the fact that early life events to influence a myriad of physical and mental health problems later in life. (22-25) We therefore build on the synthesis that, for effective and efficacious prevention of late life maladies, early life stressors need to be intervened accordingly and timely. This is because, to a sure domain, developing countries, especially Africa – and in particular Tanzania have been shown to undergo a more rapid demographic transition. (26, 27) Worse still, the ever burdened early life stress will inter alia produce an exponential and probable insurmountable consequences in coming years. For effective utilisation of time factor, reliable and valid antecedents cues (e.g. inflammatory markers, birth weights) need to be realised. There is palpable evidence to show some indicators of morbidity and mortality to have limited reliability and validity dependent on the population of interest. (28) That is up and above the likely signs that shows prevalent patterns of cardiometabolic risks among African adult population, especially in Tanzania. (29) In this regard, we wish to identify the reliability and validity of factors associated with anaemia among under-fives in a typical environment of sub-Saharan Africa.

## 2. Materials and Methods

### 2.1 Study area and population

The study will be conducted in Temeke district, Dar es Salaam - Tanzania. Temeke is the largest district in Dar es Salaam in terms of both area (729 km^2^) and population size (1,368,881) as per the most recent Tanzania housing and population census and Temeke municipal profile. [30] Dar es Salaam is a typical cosmopolitan area of sub-Saharan Africa, being rich in different demographic patterns resident in sub-Saharan Africa forms a representative population base of Africa. There are rich mix of Bantus, nilotics, Afro-Arabic as well as other Afro-Asiatic, Afro-caucasian and Indo-caucasian group members in its population base. Besides, Dar es Salaam is among the fastest growing cities in Africa and the whole world, projected to be the 10th largest city on earth in terms of population base come 2050 and rising to be the 2nd largest city on earth come 2100. [30]

### 2.2 Study design

A cross-sectional hospital-based study

### 2.3 Study duration

July – December 2022

### 2.4 Sampling

All children aged 2-59 months residing in Temeke municipality and seen at either Mbagala Rangi Tatu district hospital or Temeke Regional Referral Hospital during the study period will be eligible to participate into the study, provided that they have not received blood transfusion in the past 120 days to the recruitment date. Cochran formula will be used to determine the minimum sample size. [31]

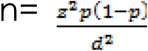

where by Z – Z-score (at α-level of 5%)

P – Proportion (prevalence) estimate from a population

D – Error term/limit of precision

In order to account for some extraneous activities, an additional 10% of the minimum sample size value will be recruited in order to account for missing data as well as refusals to participate into the study. Thus, the total sample size to be included into this study will include at least an additional 10% of the required minimum sample size.

### 2.5 Data collection tool

The study will depend on a self-administered questionnaire with five distinct parts. **Part A** of the questionnaire will consist of baseline-demographic data of the child and parents/guardians. **Part B** will consists of symptoms of anaemia including (but not limited to) fever, general body weakness, yellowish staining of the eyes, paleness of hand and feet, anorexia, cough, pica, and swelling of the body. **Part C** will constitute physical signs of anaemia, including (but not limited to) pallor, jaundice, hepatomegaly, splenomegaly, Edema, lymphadenopathy and the sign of congestive heart failure.

**Part D** will consist of anthropometric measurements of weight, height, and length. **Part E** will consist of lab parameters from a sterile collection of samples using 70% ethanol, a sterile needle, and a 5cc syringe, and placing them in a vacutainer test tube for a full blood count.

The questionnaire will be developed in English then will be translated into the Swahili language and before data collection; the pretest will be done to assess reliability and validity.

### 2.6 Data collection method

Data collection will be performed primarily by a principal investigator and two research assistants. One of the research assistants is a principal clinical officer who works in pediatric outpatient clinics at Mbagala Rangi tatu district hospital. He will be trained on Good Clinical Practice (GCP) prior to undertaking the functions.

Immediately following written informed consent from proxies (parents/guardians) data will be collected by interviewing mothers/guardians of children aged 2 to 59 months, attending outpatient clinics at either Temeke Regional Referral Hospital or Mbagala Rangi tatu district hospital using the pre-designed and pre-tested structured questionnaires, with 2-3mls of blood will be drawn from the left antecubital fossa or femoral area of the left foot of each child.

Children to be recruited will be identified in outpatient pediatric clinics at the two health facilities and after written informed consent from proxies (parents/guardians). Thereafter, the Principal Investigator and research assistants will use a pre-designed and pre-tested structured questionnaire to collect data and a complete physical examination will be performed. The axillary body temperature of each child will be measured using a digital thermometer and fever will be defined as a temperature ≥ 37.5 °C. Anthropometric measurements will include height, length, and weight and measured using a Stadiometer and weighing machine (Momerta co.Ltd Hungary). Anthrpometric measurement will be intepreted as per WHO standards with under-nutrition indices which comprised weight-for-height / length (WH/L) standard deviation (SD) scores (z-scores) to be computed based on the World Health Organization, questionnaires. The questionnaire specifically comprises of the three initials of the name, child age in terms of months, history of chronic illness and sign and symptoms like fever, general body malaise, fatigue, yellowish discoloration of eyes and heart palpitation, paleness, jaundice, lymphadenopathy, hepatomegaly. Laboratory investigations to determine hemoglobin level and red blood cell indices will be done using automated hematology analyzer, DYMIND co. limited model DH 76 (Abbott Laboratories, USA)

PI and research assistants will explain the procedure of sample collection to the mother/caregiver. Then, written informed consent will be sought from proxies to collect a blood sample from each participant. Applied tourniquet proximal to the site of venipuncture, using an aseptic technique after disinfection with cotton immersed in 70% ethanol, 2-3 milliliters of venous blood will be collected either from ante-cubital fossa or proximal femoral vein by using a 2-5 cc syringe and will be collected in EDTA vacutainer that will be stored in cold box container and will be transported from Mbagala Rangi tatu to TRRH by motorcycle for full blood picture analysis. PI and two research assistants (one is a laboratory technologist with a degree holder in medical laborateory sciences and the second one is principle clinical officer) will complete this task. PI, other investigators and laboratory technologist will be responsible to train one research assistant on how to collect samples by using standard operating procedures.

A laboratory technologist at TemekeRegional Referral Hospital will be tasked to analyze all blood samples collected and stored in ice packs containers.

### 2.7 Data analysis

Immediately following data collection, all data in questionnaire will be double entered into a pre-designed SPSS template, cleaned (checking for any incomplete data, errors in data entry) and later stored in the laptop computer of the Principal Investigator until analysis time.

Data analysis will commence initially with data exploration with summarisation of important variables of interest. Specifically, continuous data will be summarised using median (with inter-quartile range) and categorical data will be summarised using frequency (and proportion by %) Graphical tools will also be employed where by important correlations as well as +/- outliers will be assessed. Besides, univariate and bivariate statistics will be computed for all clinically relevant data.

A complete blood count is a medical laboratory finding that shows the number of white blood cells, red blood cells, and platelets in the blood. Hemoglobin concentration, hematocrit, and red blood cells morphology.

Anaemia will be reported based on hemoglobin level and red blood cell indices; and anaemia will be defined based on WHO criteria, with hemoglobin levels less than 11 g/dl being mild, less than 10 g/dl being moderate, and less than 7 g/dl being severe; and morphology of anaemia will be reported based on red blood cell indices, with mean corpuscular volume of 80 -100fl being normocytic anaemia, less than 80fl is microcytic anaemia and more than 100fl is macrocytic anaemia, mean cell hemoglobin concentration is classified as normochromic, hypochromic, or hyperchromic. (32)

For those who will be found to have severe anaemia less than 7g/dl and sepsis will be admitted for further evaluation and management, including blood transfusion and will be follow the routine procedure for health facility but will be given priority as severe anaemia and sepsis are life threatening conditions if left untreated a child will end up with complications

#### 2.7.1 Dependent variables

The severity of anemia, morphological classification, hemoglobin level, and red blood cell indices like MCV, MCHC, MCH, platelets

#### 2.7.2 Independent variables

Age, sex, residence, child anthropometric measurement (weight for height/length), fever, loss of appetite, paleness, jaundice, lymphadenopathy, hepatomegaly, and splenomegaly.

### 2.8 Ethical considerations

Ethical clearance to conduct this study has been obtained from the Institutional Research Committee of the Hubert Kairuki Memorial University with reference number HKMU/IREC/27.10/150 dated 6th of June 2022. Permission to undertake the study at the study sites will be sought from the administrations at TRRH and Mbagala Rangi tatu (via the office of the municipal director at Temeke municipal council). Parents/guardians will be approached, informed about the purpose of the study and voluntary asked for their willingness to participate in the study. Specifically, each parent will be informed about the title and objectives of the study. Besides, they will be informed of the risks and benefits associated with involvement in the study, including minimal injury during sample collection (like pain, swelling, and minor bleeding) and how they will be dealt with (e.g. bleeding solved by applying pack pressure compression at the site of bleeding for some few seconds) To prevent infection while taking sample from patient cotton immersed in 70% of ethanol and sterile disposable syringe will be used. Confidentiality will be maintained by the principal investigator and research assistant. All research assistants will undergo Good Clinical Practice (GCP) training prior to recruitment into the study as personnel. All patients’ details will be private and unique by assigning codes to each participant except for the laboratory requisition form. Parent/guardian should consent for children, in this study, participants will have the right to refuse/ withdraw from the study and it will not hinder them to receive their medical care.

## 2.9 Study limitations

### Failure to show temporal association between variables

The study has been designed in a cross-sectional fashion, and hence it will not be possible to account for any (if present) temporal association between studied variables.

### Respondent with incomplete data

There is a possibility that part of participants data will be missing during collection time (e.g. missing information on lab findings) since the study has been designed in a cross-sectional fashion.

### Record bias

It is also likely that part of the collected information will be erroneous or biased (positively or negatively) altogether by the time of collection from achieved data (e.g. laboratory findings and/or vaccination cards)

## Data Availability

No data is available for now. It is a protocol

## References

1. Stevens G, Finucane M, De-Regil L, Paciorek C, Flaxman S, Branca F, et al. Global, regional and national trends in haemoglobin concentration and prevalence of total and severe anaemia in children and pregnant and non-pregnant women for 1995-2011: a systematic analysis of population-representative data. Lancet Global Health 2013; 1: e16–e25.

2. Sarna A, Porwal A, Ramesh S, Agrawal P, Acharya R, Johnston R, et al. Characterisation of the types of anaemia prevalent among children and adolescents aged 1-19 years in India: a population based study. Lancet Child & Adolescent Health 2020; 4(7): P515–P525.

3. Dos Santos R, Gonzalez E, Albuquerque E, Arruda I, Diniz A, Figueroa J, et al. Prevalence of anaemia in under-five years old children in a children’s hospital in Recife, Brazil. Rev Bras Hematol Hemoter. 2011; 33(2): 100–104.

4. Sun J, Wu H, Zhao M, Magnussen C. and Xi B. Prevalence and changes of anaemia among young children and women in 47-low and middle-income countries, 2000-2018. Eclinical Medicine 2021; 41: 1–9.

5. Kejo D, Petrucka P, Martin H, Kimanya M. and Mosha T. Prevalence and predictors of anemia among children under 5 years of age in Arusha district, Tanzania. Pediatric Health, Medicine & Therap. 2018; 9; 9–15.

6. Schellenberg D, Armstrong-Schellenberg J, Mushi A, Savigny D, Mgalula L, Mbuya C. and Victoria C. The silent burden of anaemia in Tanzanian children: a community-based study. Bull World Health Org. 2003; 81(8): 581–590.

7. Premji Z, Hamisi Y, Shiff C, Minjas J, Lubega P. and Makwaya C. Anaemia and plasmodium falciparum infections among young children in holoendemic area, Bagamoyo, Tanzania. Acta Tropica 1995; 59: 55–64.

8. Abdulla S, Schellenberg J, Nathan R, Mukasa O, Marchant T, Smith T, et al. Impact of malaria morbidity of a programme supplying insecticide treated nets in children aged under 2-years in Tanzania: community cross-sectional study. BMJ 2001; 322: 270–273.

9. Barker D, Winter P, Osmond C, Margetts B. and Simmonds S. Weight in infancy and deaths from ischaemic disease. Lancet 1989; 2: 577–580.

10. Preston S, Hill M. and Drevenstedt G. Childhood conditions that predict survival to advanced ages among African-Americans. SOSMED 1998; 47(9): 1231–1246.

11. McDade T, Beck M, Kuzawa C. and Adair L. Prenatal undernutrition, postanatal environment and antibody response to vaccinations in adolescence. Am. J Clin. Nutr. 2001; 74: 543–548.

12. Plan N, Reproductive FOR, Health A. National Plan for Reproductive, Maternal, Newborn, Child and Adolescent Health & Nutrition (2021/2022 – 2025/2026). One Plan I. 2022. 10.

13. World Bank. Public Health at a glance – Anemia. Available from http://web.worldbank.org/archive/website01213/WEB/0 [Accessed on 1 October 2022] CO-50.HTM

14. World Health Organisation. Anaemia Policy Brief. Available from WHO_NMH_NHD_14.4_eng.pdf. [Accessed on 1 October 2022]

15. Stevens G, Finucane M, De-Regil L, Paciorek C, Flaxman S, Branca F, et al. Global, regional and national trends in haemoglobin concentration and prevalence of total and severe anaemia in children and pregnant and non-pregnant women for 1995-2011: a systematic analysis of population-representative data. Lancet Global Health 2013; 1(1): E16–E25.

16. Babu DUP, Prasad Dbvs, Reddy ES, Manasa R V. A Cross Sectional Study on Morphological Pattern of Anemia. IOSR J Dent Med Sci [Internet]. 2016;15(08):68–71. Available from: 10.9790/0853-1508086871.

17. World Health Organisation. Haemoglobin concentrations for the diagnosis of anaemia and assessment of severity. Geneva, Switz World Heal Organ [Internet]. 2011;1–6. Available from: http://scholar.google.com/scholar?hl=en&btnG=Search&q=intitle:Haemoglobin+concentrations+for+the+diagnosis+of+anaemia+and+assessment+of+severity#1

18. Mboya IB, Mamseri R, Leyaro BJ, George J, Msuya SE, Mgongo M. Prevalence and factors associated with anemia among children under five years of age in Rombo district, Kilimanjaro region, Northern Tanzania. F1000Research [Internet]. 2020 Sep 7;9:1102. Available from: https://f1000research.com/articles/9-1102/v1.

19. Temeke council. Health Profile - Temeke Municipal Council. 2017; Available from: http://www.temekemc.go.tz/storage/app/uploads/public/59d/ba8/62e/59dba862ed84d383294414.pdf

20. Miller G, Chen E, Fok A, Walker H, Lim A, Nicholls E, et al. Low early-life social class leaves a biological residue manifested by decreased glucocorticoid and increased proinflammatory signalling. Proceedings of the National Academy of Sciences of the United States of America 2009; 106(34): 14716–14721.

21. Kananen L, Surraka I, Pirkola S, Suvisaari J, Lonnqvist J, Peltonen L, et al. Childhood adversities are associated with shorter telomere length at adult age both in individuals with an anxiety disorder and controls. PLoS ONE 2010; 5(5): e10826.

22. Taylor S. Mechanisms linking early life stress to adult health outcomes. PNAS 2010; 107(19): 8507–8512.

23. Barker D, Osmond C, Winter P, Margetts B. and Simmonds S. Weight in infancy and death from ischaemic heart disease. Lancet 1989; 334(8663): 577–580.

24. Resolution WHA 65.6. Comprehensive implementation plan on maternal, infant and young child nutrition. In: Sixty-fifth World Health Assembly Geneva, 21-26 May 2012. Resolutions and decisions, annexes. Geneva: World Health Organisation 2012; 12–13. Available from http://apps.who.int/iris/bitstream/handle/10665/113048/WHO_NMH_NHD_14.1_eng.pdf?sequence=1 [Accessed on 1 August 2022]

25. World Health Organisation. Global targets 2025. To improve maternal, infant and young child nutrition. Available from http://www.who.int/nutrition/topics/nutrition_globaltargets2025/en/ [Accessed on 1 August 2022]

26. Leshabari K. Demographic transition in sub-Saharan Africa: from grassroots to ivory towers. In: Klimczuk (ed) Demographic analysis: selected concepts and analysis. IntechOpen: London, 2021. Available from https://www.intechopen.com/chapters/77806 [Accessed on 1 August 2022]

27. Leshabari K, Biswas A, Gebuis E, Leshabari S. and Ohnishi M. Challenges in morbidity and mortality statistics of the elderly population in Tanzania: a call to action. Quality in Ageing & Older Adults 2017; 18(3): 171–174.

28. Leshabari K. Reliability and validity of the clinicopathological features associated with frailty syndrome in elderly population. In Palermo S. (ed) Frailty in the elderly: understanding and managing complexity. IntechOpen: London, 2021. Available from https://www.intechopen.com/chapters/73143 [Accessed on 7 August 2022]

29. Leshabari K, Ndonde W, Mbega S. and Mbululo L. Predictors of cardiometabolic risks among typical African elderly population: analysis from Dar es Salaam, Tanzania. Diabetes Technology & Therapeutics 2020; 22: A214.

30. Dar es Salaam population 2022. Available from https://worldpopulationreview.com/world-cities/dar-es-salaam-population [Accessed on 7 August 2022]

31. Cochran W. Sampling techniques. John Wiley & Sons, 1977.

32. Cells RB. Complete Blood Count Normal Pediatric Values Complete Blood Count Normal Pediatric Values Differential White Blood Cell Count Normal Pediatric Values. :22.

